# A Distal–Proximal Gradient of Hebbian Plasticity in the Human Arm

**DOI:** 10.64898/2026.09.23.26363732

**Authors:** Minkyu Lee, Matthew T. Farley, Bing Chen, Sina Sangari, Monica A. Perez

**Author notes:** **Corresponding author:** Monica A. Perez, PT, PhD, Scientific Chair, Arms + Hands Lab at Shirley Ryan AbilityLab, Professor, Department of Physical Medicine and Rehabilitation at Northwestern University Research Scientist, Hines VA Medical Center, office.

## Abstract

Corticospinal–motoneuronal connections are more prominent in distal than proximal upper-limb muscles, but whether this anatomical specialization influences the capacity for activity-dependent plasticity remains unknown. Here, we tested whether Hebbian stimulation targeting corticospinal–motoneuronal connections, induces muscle-specific plasticity in proximal and distal arm muscles in healthy humans. We applied a spike-timing-dependent stimulation protocol comprising 180 paired stimuli, with transcranial magnetic stimulation timed to evoke corticospinal volleys 1–2 ms before motoneurons were antidromically activated by electrical stimulation of the brachial plexus for the biceps brachii or the ulnar nerve for the first dorsal interosseous (FDI). Corticospinal excitability was assessed by measuring motor-evoked potentials (MEPs) before and 30 min after stimulation. Hebbian stimulation increased MEP amplitudes in both muscles, but facilitation was greater in the FDI than in the biceps. Because resting motor threshold was higher for the biceps than for the FDI, in a control experiment we matched stimulation intensities between muscles. Under these conditions, Hebbian stimulation continued producing greater facilitation in the FDI than in the biceps, indicating that the difference was not attributable to stimulation intensity. Moreover, changes in MEP amplitude were associated with maximal MEP size in the FDI but not in the biceps, whereas no muscle-specific association was observed with peripheral responses (M-max), supporting a stronger relationship between corticospinal drive and Hebbian plasticity in the distal than in the proximal arm muscle. Together, these findings identify a distal-to-proximal gradient in Hebbian plasticity across arm muscles, which may contribute to optimizing targeted neuromodulation approaches for upper-limb rehabilitation.

**Highlights:**

- Plasticity in distal and proximal upper-limb muscles remains poorly understood
- Ability to induce Hebbian plasticity was greater in a hand than in proximal arm muscle
- A distal-to-proximal gradient in Hebbian plasticity may inform targeted rehabilitation

## Introduction

Anatomical and electrophysiological studies have demonstrated that corticospinal projections differ between proximal and distal upper-limb muscles. Specifically, in non-human primates, distal muscles of the hand receive denser direct monosynaptic corticospinal– motoneuronal projections than proximal arm muscles, which receive less prominent monosynaptic corticospinal input [1, 2]. Consistent with these observations, studies in humans using peristimulus time histograms (PSTHs) of single motor unit discharges have shown stronger corticospinal facilitation of finger motoneurons than proximal arm motoneurons [3]. Furthermore, motor-evoked potentials (MEPs) elicited by transcranial magnetic stimulation are larger and have a lower threshold in finger muscles than in proximal arm muscles [4]. Whether these differences affect the ability to induce activity-dependent plasticity in corticospinal projections to proximal and distal upper-limb muscles remains unknown.

Hebbian stimulation, based on the principles of spike-timing-dependent plasticity (STDP), provides a powerful approach to investigate muscle-specific differences in corticospinal plasticity. By precisely timing cortical and peripheral inputs, this intervention aims to strengthen corticospinal–motoneuronal connections through mechanisms resembling long-term potentiation [5–9]. Hebbian stimulation enhances corticospinal plasticity in both healthy individuals and people with spinal cord injury [10–13]. Because STDP depends on the precise timing of presynaptic and postsynaptic activity, its efficacy may be influenced by the strength and organization of corticospinal connectivity. Consistent with this view, Hebbian-induced facilitation is greater when stimulation is delivered during low-level voluntary contraction than at rest, when descending drive to motoneurons is more actively engaged [14]. Thus, muscles with stronger direct corticospinal connections may exhibit greater Hebbian-induced plasticity. Indeed, evidence from previous studies supports the existence of muscle-specific differences in other forms of plasticity. For example, pairing TMS with motor point stimulation produced the largest increases in corticospinal excitability in intrinsic hand muscles, smaller increases in the flexor digitorum superficialis, and no detectable changes in the extensor digitorum communis [15]. In addition, Hebbian stimulation produces greater increases in corticospinal excitability in elbow flexor than extensor muscles, supporting the possibility of inducing muscle-specific plasticity [16]. We hypothesized that the first dorsal interosseous (FDI), a distal muscle with stronger monosynaptic corticospinal input, would exhibit greater increases in corticospinal excitability after Hebbian stimulation than the biceps, a proximal muscle with comparatively weaker direct corticospinal connectivity. Understanding muscle-specific differences in corticospinal plasticity may help optimize neuromodulation strategies for targeted rehabilitation.

To test this hypothesis, we compared the effects of Hebbian stimulation on MEP amplitude in the FDI and biceps brachii in separate sessions. Hebbian stimulation consisted of 180 precisely timed paired stimuli designed to induce STDP-like plasticity by delivering corticospinal volleys 1–2 ms before antidromic motoneuron activation via electrical stimulation of the brachial plexus (biceps) or ulnar nerve (FDI). MEP responses were assessed before and for up to 30 min after stimulation.

## Materials & Methods

### Subjects

Sixteen neurologically intact individuals (mean age, 31±10 years; 8 females and 8 males) participated in the study. All participants provided written informed consent before experimental procedures, which were approved by the local ethics committee at Northwestern University (STU00210458) and conducted in accordance with the Declaration of Helsinki. Participants had no history of neurological or orthopedic injuries affecting the upper limb, and the dominant arm was tested in all experiments.

### Electromyographic (EMG) recordings

EMG activity was recorded from the FDI and biceps brachii using bipolar surface electrodes (Ag–AgCl, 10 mm diameter; 1 cm interelectrode distance) placed over the muscle belly. Signals were amplified, band-pass filtered (30–2000 Hz), and sampled at 4 kHz for offline analysis (CED 1401 with Signal software; Cambridge Electronic Design, Cambridge, UK).

### Experimental setup

During testing of the FDI muscle, participants were seated in an armchair with both arms relaxed, elbows flexed to 90°, and forearms pronated. When testing the biceps brachii, the tested arm was supported on a custom platform with the shoulder and elbow positioned at 90° flexion. The wrist and forearm were secured using cushioned straps to minimize movement. The testing of each muscle was conducted in different sessions separated by a few days. During Hebbian stimulation, TMS and peripheral nerve stimulation (PNS) were delivered to target corticospinal-motoneuronal connections to the FDI and biceps (Figure 1A) using 180 paired pulses at 0.2 Hz (see details below). In the main experiment, MEPs were recorded before Hebbian stimulation (Pre) and after Hebbian stimulation at 0 and 30 min post-intervention (Post0 and Post30; Figure 1B). We measured maximal MEP amplitude (MEP-max: FDI=5.6±3.4 mV, biceps=1.1±0.9 mV; p<0.001, Figure 2A), maximal motor response (M-max: FDI=17.7±6.0 mV, biceps=10.5±4.3 mV; p=0.003, Figure 2B), and resting motor threshold (RMT: FDI=53±8 % maximum stimulus intensity (MSO), biceps = 65±16 % MSO; p=0.001, Figure 2C) in both muscles.

**Figure 1.**
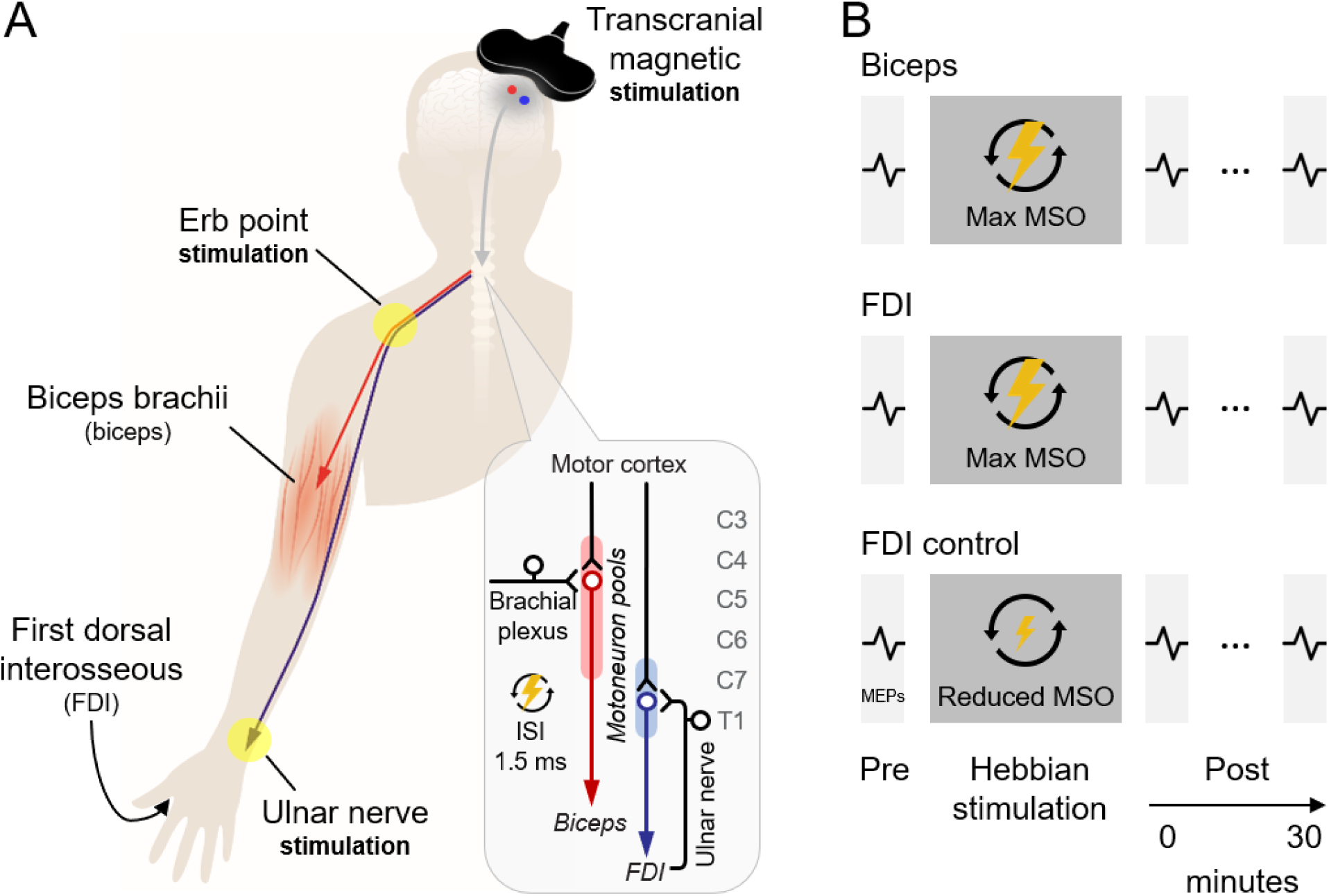
Experimental set-up and study design. (A) Illustration of the Hebbian stimulation protocol. Suprathreshold transcranial magnetic stimulation (TMS) was applied over the arm and hand representations of the primary motor cortex to activate corticospinal neurons (gray lines). Peripheral nerve stimulation (PNS) was delivered supramaximally at Erb’s point (brachial plexus) and at the ulnar nerve at the wrist. The circular callout depicts the interstimulus interval (ISI) between paired pulses: descending volleys elicited by TMS reached presynaptic terminals of corticospinal neurons 1–2 ms before antidromic volleys induced by PNS reached spinal motoneurons. (B) Participants completed three experimental sessions separated by several days. Motor evoked potentials (MEPs) in the first dorsal interosseous (FDI) and biceps muscles were recorded before and after Hebbian stimulation (Post0 and Post30). In the first session, the biceps was tested before the FDI to ensure that both Hebbian stimulation protocols could be completed in all participants. In the final session, because the resting motor threshold (RMT) was higher for the biceps than for the FDI, a control experiment was performed in which Hebbian stimulation targeting the FDI was delivered using stimulus intensities matched to the biceps RMT (FDI control).

**Figure 2.**
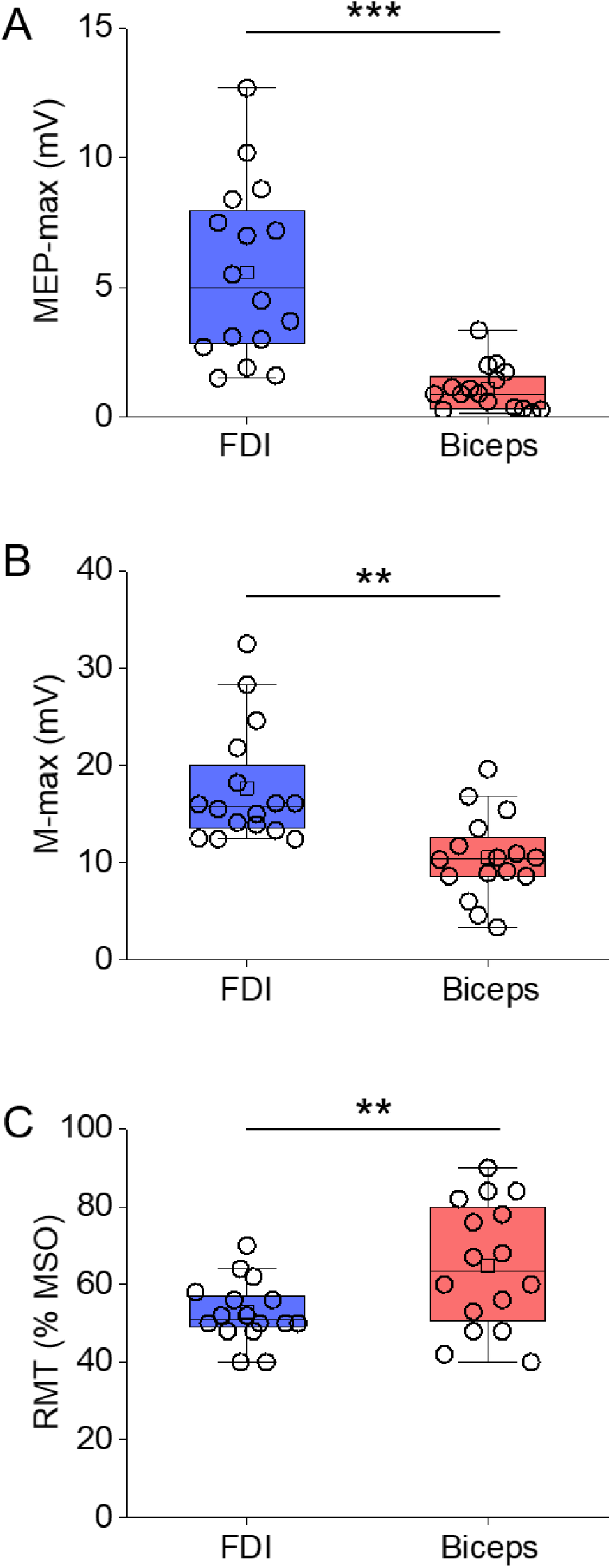
MEP-max, M-max, RMT. Group data for maximal motor-evoked potential (MEP-max) amplitude, maximal motor response (M-max) amplitude, and resting motor threshold (RMT) (up to lower) in the first dorsal interosseous (FDI) and biceps muscle. The abscissa indicates the muscle tested (FDI and biceps). The ordinate shows MEP-max amplitude (millivolts, A), M-max amplitude (millivolts, B), and RMT (maximum stimulator output, C) in the FDI and biceps. **p<0.01 and ***p<0.001.

Because the RMT was higher in the biceps compared to the FDI and, in some subjects, it is difficult to elicit a clear MEP in the biceps, we tested the biceps muscle before FDI to ensure that we were able to complete both sessions of Hebbian stimulation in all participants. Furthermore, because the RMT was higher in the biceps compared to the FDI, we completed a control experiment (referred to as ‘FDI control’, n=13) in which Hebbian stimulation targeting the FDI was applied using stimulus intensities that matched the RMT of the biceps. Thus, in this control experiment, the stimulus intensity used during Hebbian stimulation targeting the FDI was set proportionally to the intensity used for the biceps with respect to the RMT. The average intensity used during Hebbian stimulation targeting the biceps was 100 % of the MSO, which corresponded to 165±44 % of the biceps RMT. The stimulus intensity used during Hebbian stimulation targeting the FDI control was 87±14 % MSO, which corresponded to 170±47 % of the FDI RMT (p=0.447 vs. biceps).

### TMS

TMS was delivered from a Magstim 200 stimulator (Magstim company) through a figure-of-eight coil (loop diameter, 7 cm; type number SP15560) with a monophasic current waveform. TMS was applied over the hand and arm representations of the primary motor cortex, with the coil positioned at the optimal site for eliciting MEPs in the FDI and biceps. To identify the optimal scalp position for each muscle, the coil was held tangential to the scalp, with the handle angled 45° away from the midline, and moved in small steps along each motor representation. RMT for each muscle was defined as the stimulator intensity required to elicit MEPs with peak-to-peak amplitudes ≥50 µV in 5 of 10 consecutive trials at rest. Participants wore a cap with the coil position marked to ensure consistent stimulation location throughout the experiment.

### PNS

Transcutaneous electrical stimulation was applied to the ulnar nerve at the wrist for the FDI and the brachial plexus for the biceps, using a DS7R stimulator (Digitimer Ltd, UK) with a pulse width of 200 µs. For the FDI, the anode and cathode electrodes were 3 cm apart and 1 cm in diameter, with the cathode positioned proximally. For the biceps, the cathode was placed over Erb’s point and the anode over the acromion. Stimuli were delivered supramaximally at 120 % of the intensity required to elicit the M-max in the FDI and biceps. The M-max was determined by gradually increasing stimulus intensity until no further increase in M-wave amplitude was observed. M-max was measured as the peak-to-peak amplitude in millivolts (mV) of the non-rectified response.

### Hebbian stimulation

Hebbian stimulation was performed at rest, aiming to target corticospinal– motoneuronal synapses by precisely timing the interstimulus interval (ISI) between TMS and PNS. The ISI was adjusted so that descending corticospinal volleys elicited by TMS arrived at the presynaptic terminals of corticospinal neurons 1–2 ms before antidromic volleys from PNS reached the motoneuron postsynaptic terminals. TMS intensity was set to 100 % of the MSO, and PNS intensity to 120 % of the intensity that elicited the M-max for each muscle. The ISI was determined for each participant based on peripheral conduction time (PCT) and central conduction time (CCT; Figure 3). For the FDI muscle, PCT was calculated as (F-wave latency − M-max latency) × 0.5, and CCT was calculated as MEP latency − (PCT + M-max latency). For the biceps muscle, CCT was calculated as MEP latency − (C-root latency + 1.5 ms), PCT was calculated as C-root latency − M-max latency + 0.5 ms, and the ISI was calculated as CCT + 1.5 ms – PCT. TMS was used over the C5-C6 cervical spinal processes to determine the C-root latency. C-root and M-max latencies (for biceps) and F-wave (for FDI) were measured at rest using averaged waveforms, and onset latencies were identified as the first time point exceeding two standard deviations above the mean rectified pre-stimulus background activity measured over 100 milliseconds (ms) before stimulation. MEP latencies were recorded during isometric contraction at 10 % of maximum voluntary contraction (MVC), with TMS intensity set at ∼120 % of RMT. MVCs were measured for each muscle using 3–5 sec of isometric contractions: elbow flexion for the biceps and index finger abduction for the FDI. Each contraction was repeated three times with 1 min rest intervals. The maximum mean rectified EMG activity over a 1-sec epoch from each trial was calculated, and the highest value was used as the MVC reference for each muscle.

**Figure 3.**
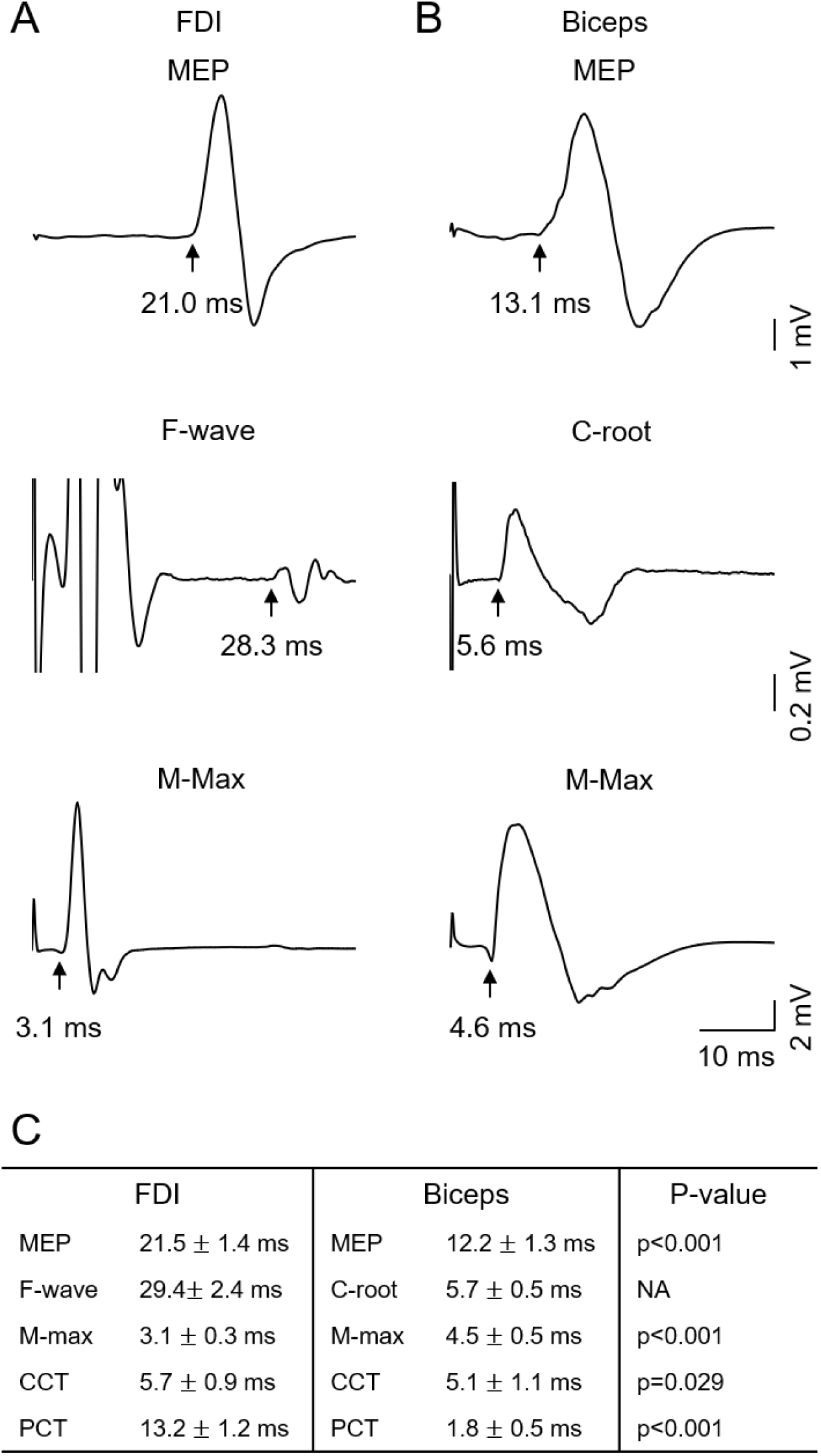
Response latencies. Raw traces showing a motor-evoked potential (MEP), an F-wave, C-root responses, and the maximal motor response (M-max) for a representative participant from (A) the FDI and (B) biceps muscles. Averaged waveforms of representative raw traces are shown. (C) MEP, F-wave, C-root, and M-max latencies were used to calculate central (CCT) and peripheral (PCT) conduction times used to estimate the time of arrival of pre- and postsynaptic volleys at cortico-motoneuronal synapses during Hebbian stimulation in each subject.

### MEPs

MEPs recorded before Hebbian stimulation were 3–5 % of M-max in amplitude in both the biceps (5.1±2.5 % of M-max) and FDI (3.9±1.4 % of M-max; p=0.122; n=16). In the FDI control experiment, MEPs recorded before Hebbian stimulation were also 3–5 % of M-max in amplitude (FDI control: 3.7±0.9 % of M-max; p=0.780 vs. FDI; n=13). Measurements were obtained immediately after Hebbian stimulation and up to 30 min thereafter. At baseline (Pre), two sets of 15 MEPs and, after Hebbian stimulation (Post), two sets of 15 MEPs were recorded at rest. The peak-to-peak amplitude of each MEP was measured, and the average amplitude across trials was calculated for each time point. Trials with background EMG activity ≥25 µV were excluded from analysis (9 % of trials were excluded). For analysis, MEP amplitudes were normalized to baseline (Pre), expressed as percent change from baseline, and compared across post-stimulation time points (Post0 and Post30).

### Data Analysis

Mauchly’s test was used for testing sphericity. When the assumption of sphericity was violated, the Greenhouse–Geisser was applied. One-way repeated measures ANOVA was used to examine the effect of TIME (Pre, Post0, and Post30) on normalized MEP amplitude in both muscles. A two-way repeated measures ANOVA was used to examine the effect of TIME, MUSCLE (biceps, FDI, FDI control), and their interaction on normalized MEP amplitude. Paired t-tests were used to compare MEP-max, RMT, and M-max between biceps and FDI. Bonferroni corrections were used as *post hoc* tests to assess for significant comparisons. Pairwise comparisons between the biceps, FDI, and FDI control conditions were conducted at each post-stimulation time point (Post0 and Post30) using Fisher’s least significant difference (LSD) test. Statistical analysis was conducted using SPSS (IBM Corp. Released 2019. IBM SPSS Statistics for Windows, Version 26.0. Armonk, NY: IBM Corp), and the significance was set at p<0.05. ANOVA effect size is reported for significant results with partial η^2^. Group data are presented as means ± SDs in the text.

## Results

### MEPs

Figure 4A illustrates raw MEP traces from the FDI of a representative participant. Repeated-measures ANOVA revealed significant effects of TIME (F_2,30_=13.696, p<0.001, partial η²=0.477) on normalized MEP amplitude. *Post hoc* analysis showed that following Hebbian stimulation, MEP amplitudes increased, and this enhancement persisted for up to 30 min. We found that MEP amplitudes were increased compared with their baseline at Post0 (233±132 %; p=0.004) and Post30 (239±144 %; p=0.005), with an overall average increase of 236±134 % across time points. Individual data showed that 15 of 16 participants exhibited increased MEP amplitudes compared to their baseline at Post0 and 15 of 16 participants increased MEP amplitudes compared to their baseline at Post30 (Figure 4C).

**Figure 4.**
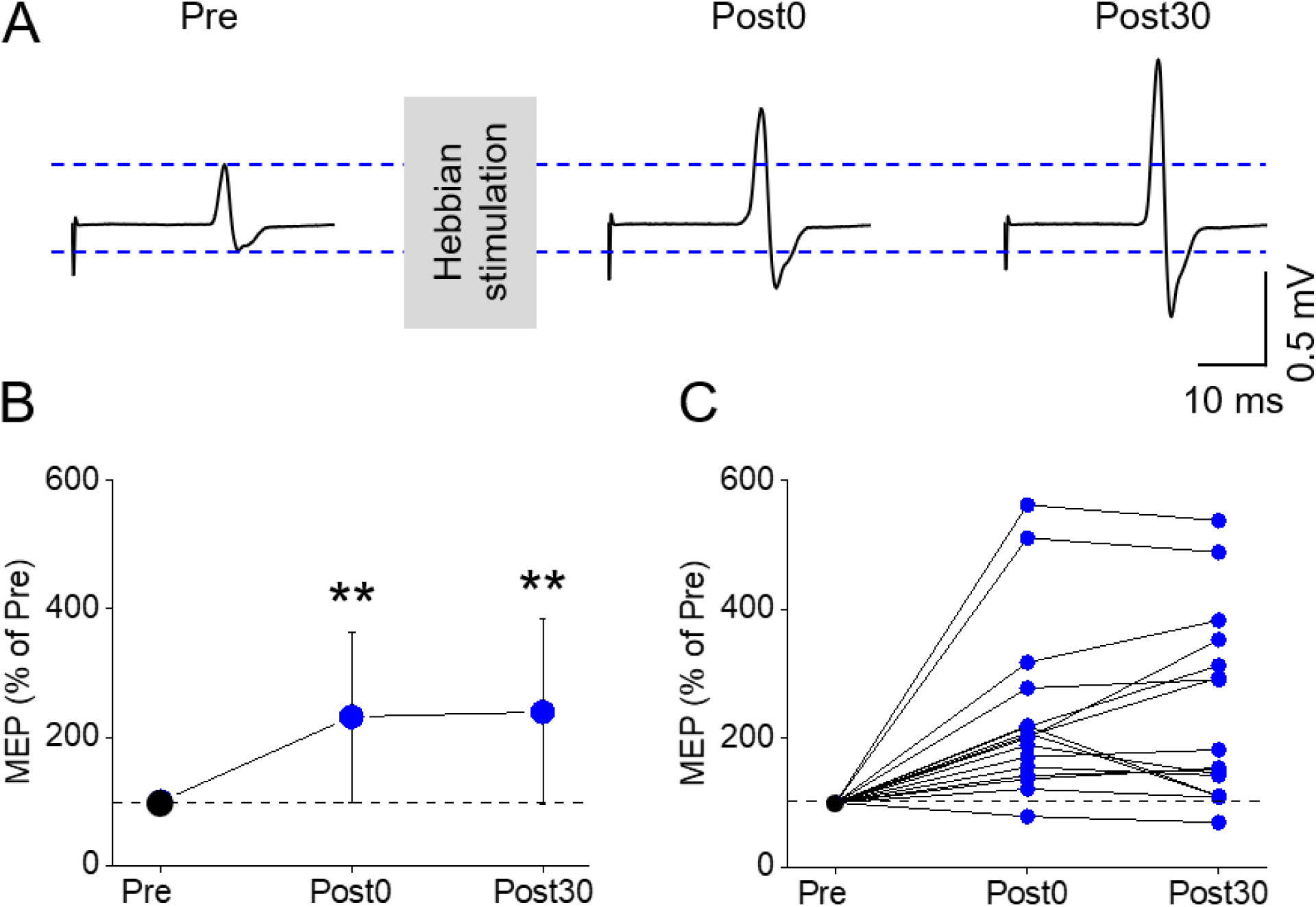
MEPs in the FDI muscle. (A) Representative motor-evoked potential (MEP) traces elicited by transcranial magnetic stimulation in the first dorsal interosseous (FDI) muscle before (Pre) and after Hebbian stimulation (Post0 and Post30). (B) Group data showing MEP amplitudes expressed as a percentage of the pre-stimulation value. MEPs were measured before (Pre), immediately after (Post0), and 30 min after (Post30) Hebbian stimulation. (C) Blue circles represent individual participant data showing average MEP amplitudes relative to Pre at each time point after Hebbian stimulation. The horizontal dashed line in panels B and C indicates the baseline (Pre) MEP amplitude (100 %). Error bars represent standard deviation. **p<0.01.

Figure 5A illustrates raw MEP traces from the biceps of a representative participant. Repeated-measures ANOVA revealed significant effects of TIME (F_2,30_=4.549, p=0.019, partial η²=0.233) on normalized MEP amplitude. *Post hoc* analysis showed that following Hebbian stimulation, MEP amplitudes increased compared to baseline at Post0 (125±34 %; p=0.030) and Post30 (125±37 %; p=0.045), with an overall average increase of 125±28 % across time points. Individual data showed that 12 of 16 participants exhibited an increase in average MEP amplitudes compared to their baseline at Post0 and 13 of 16 participants increased MEP amplitudes compared to their baseline at Post30 (Figure 5C).

**Figure 5.**
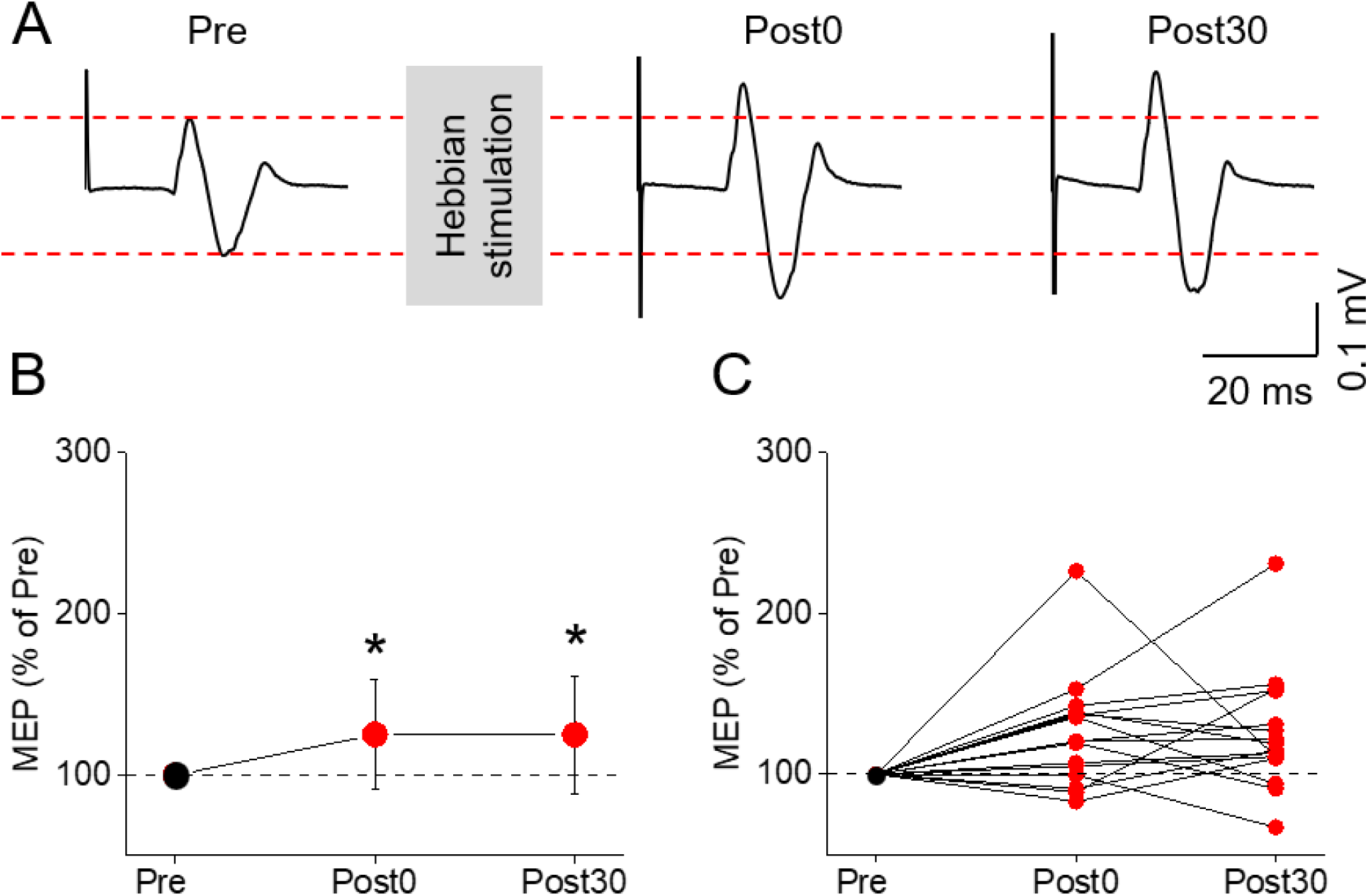
MEPs in the biceps muscle. (A) Representative motor-evoked potential (MEP) traces elicited by transcranial magnetic stimulation in the biceps muscle before (Pre) and after Hebbian stimulation (Post0 and Post30). (B) Group data showing MEP amplitudes expressed as a percentage of the pre-stimulation value. MEPs were measured before (Pre), immediately after (Post0), and 30 min after (Post30) Hebbian stimulation. (C) Red circles represent individual participant data showing average MEP amplitudes relative to Pre at each time point after Hebbian stimulation. The horizontal dashed line in panels B and C indicates the baseline (Pre) MEP amplitude (100%). Error bars represent standard deviation. *p<0.05.

### MEPs in the FDI vs. the biceps brachii

Figure 6A illustrates raw MEP traces from the FDI (top traces), biceps (middle traces), and FDI control (bottom traces) of a representative participant. MEP amplitude increased to a similar extent in the FDI and FDI control conditions, with both showing greater increases than the biceps following Hebbian stimulation.

**Figure 6.**
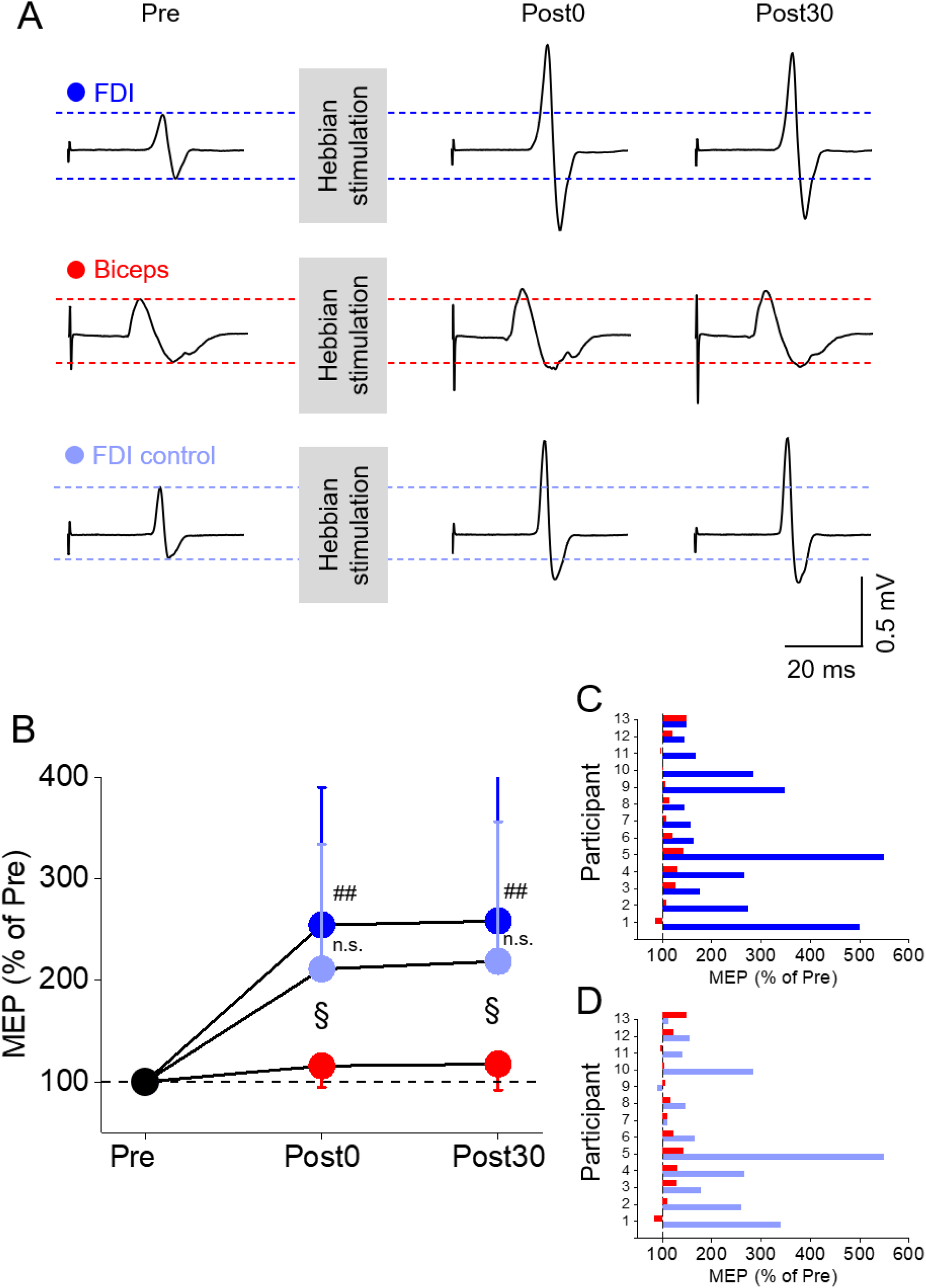
MEPs in the FDI and biceps muscle. (A) Representative motor-evoked potential (MEP) traces elicited by transcranial magnetic stimulation (TMS) before (Pre) and after Hebbian stimulation (Post0 and Post30) in the FDI at 100 % maximum stimulator output (MSO), the biceps at 100 % MSO, and the FDI control condition, in which the TMS intensity during Hebbian stimulation was matched to the resting motor threshold of the biceps. (B) Group data showing MEP amplitudes expressed as a percentage of the pre-stimulation value. MEPs were measured before (Pre), immediately after (Post0), and 30 min after (Post30) Hebbian stimulation in the FDI (blue circles), FDI control (light blue circles), and biceps (red circles). Error bars represent standard deviation. (C) Individual participant data showing average MEP amplitudes relative to Pre at each time point after Hebbian stimulation for the FDI and biceps. (D) Individual participant data showing average MEP amplitudes relative to Pre at each time point after Hebbian stimulation for the FDI control and biceps. ^##^p<0.01 for the comparison between FDI and biceps; ^§^p < 0.05 for the comparison between FDI control and biceps. n.s. (not significant) for the comparison between FDI and FDI control.

Repeated-measures ANOVA (Figure 6B) revealed significant main effects of TIME (F₂,₂₄=15.049, p<0.001, partial η²=0.556) and MUSCLE (F₂,₂₄=9.657, p=0.001, partial η²=0.446), as well as a significant TIME × MUSCLE interaction (F₄,₄₈=7.202, p<0.001, partial η²=0.375) on normalized MEP amplitude. *Post hoc* analysis showed greater facilitation in the FDI than in the biceps at both Post0 (254±136 % vs. 116±21 %, respectively; p=0.003) and Post30 (258±147 % vs. 118±26 %, respectively; p=0.007). Across Post0 and Post30, the average facilitation was 256±137 % in the FDI and 117±19 % in the biceps. Individual data (FDI vs. biceps) are shown in Figure 6C. At the individual level, all participants exhibited greater MEP facilitation in the FDI than in the biceps at Post0, and 10 participants exhibited greater facilitation at Post30 following Hebbian stimulation. Individual data (FDI control vs. biceps) are shown in Figure 6D. At the individual level, 10 participants exhibited greater MEP facilitation in the FDI than in the biceps at Post0, and 10 participants exhibited greater facilitation at Post30 following Hebbian stimulation.

*Post hoc* analysis also showed greater facilitation in the FDI control than in the biceps at Post0 (211±123 % vs. 116±21 %, respectively; p=0.015) and Post30 (219±138 % vs. 118±26 %, respectively; p=0.023). In contrast, facilitation did not differ between the FDI and FDI control conditions at either Post0 (p=0.121) or Post30 (p=0.114).

### Correlation analysis

Figure 7 illustrates the correlations between MEP-max and M-max with the average post-intervention MEP facilitation, calculated as the average of Post0 and Post30, for both muscles. In the FDI, MEP-max was positively correlated with average MEP facilitation (r=0.694, p=0.003; Figure 7A), and in the FDI Control (r=0.783, p=0.002). In contrast, MEP-max was not significantly correlated with average MEP facilitation in the biceps (r=0.341, p=0.196; Figure 7B). In terms of the periphery, FDI M-max was positively correlated with the average post-intervention MEP response in both the FDI (r=0.712, p=0.002; Figure 7C) and FDI control (r=0.747, p=0.003) conditions. Similarly, biceps M-max was positively correlated with the average post-intervention MEP response (r=0.689, p=0.003; Figure 7D).

**Figure 7.**
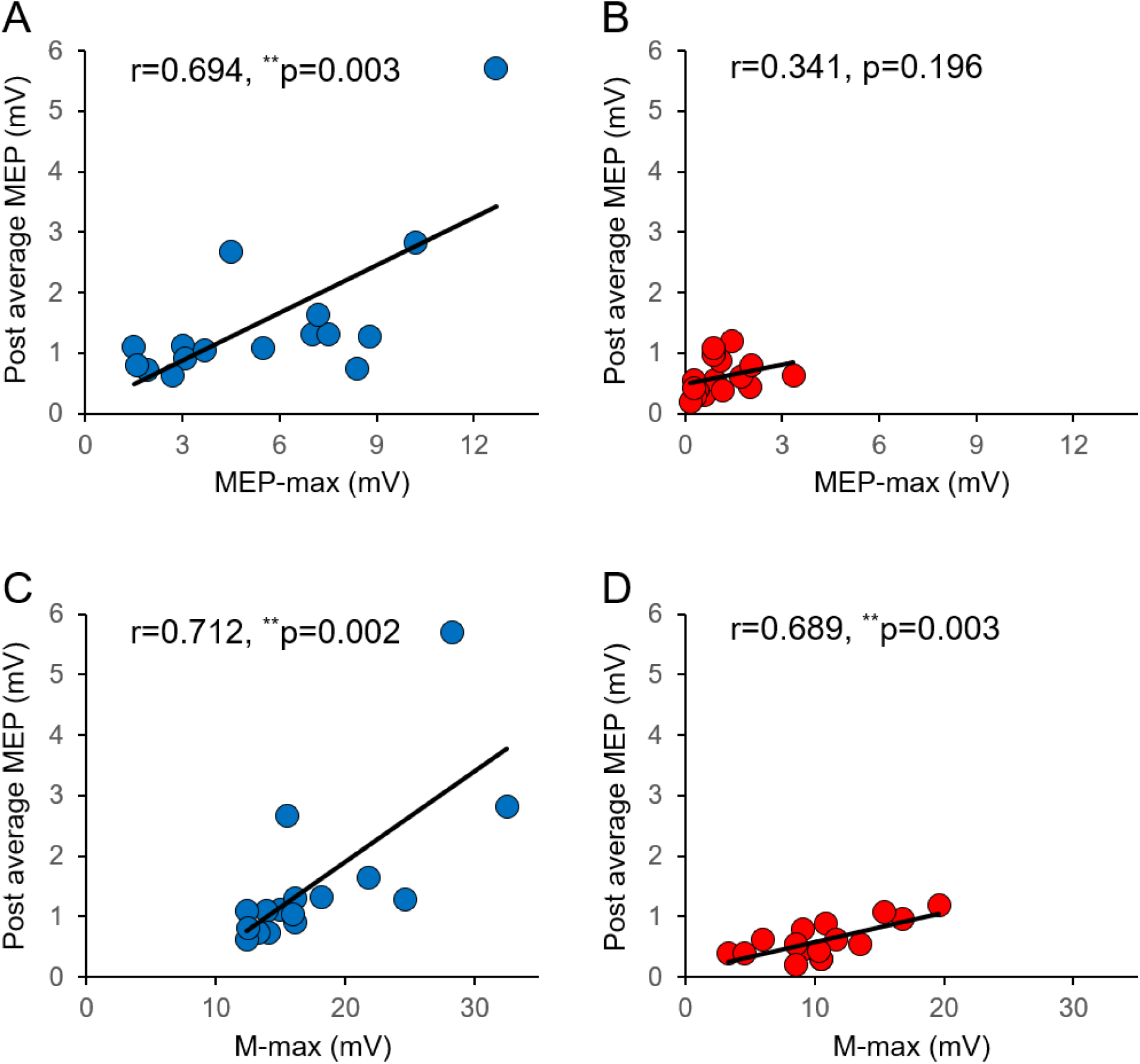
Correlations. Individual data for the FDI (blue) and biceps (red) are shown. In the upper graphs, the abscissa indicates the maximal motor-evoked potential (MEP-max) amplitude in the FDI (A) and biceps (B), and the ordinate shows the average change in MEP amplitude after Hebbian stimulation targeting each muscle. In the lower graphs, the abscissa indicates the maximal motor response (M-max) amplitude in the FDI (C) and biceps (D) muscles, and the ordinate shows the average change in MEP amplitude after Hebbian stimulation targeting each muscle. In all graphs, the solid line represents the Pearson correlation calculated across all data points from all participants tested. Notably, changes in MEP amplitude after Hebbian stimulation were associated with MEP-max amplitude in the FDI but not in the biceps, whereas no muscle-specific association was observed with the M-max. **p<0.01.

## Discussion

Our findings demonstrate, for the first time, a distal-to-proximal gradient in Hebbian plasticity between proximal and distal arm muscles. Hebbian stimulation targeting corticospinal– motoneuronal connections increased MEP amplitude in both the FDI and biceps, but the magnitude of facilitation was significantly greater in the distal FDI. Note that the resting motor threshold was higher for the biceps brachii than for the FDI. Therefore, in the FDI control experiment, we tested whether this difference could be attributed to differences in stimulation intensity. Importantly, the greater facilitation in the FDI persisted when stimulation intensities were matched between muscles, indicating that it cannot be explained simply by differences in stimulation thresholds. Notably, changes in MEP amplitude were associated with maximal MEP size in the FDI but not in the biceps, whereas no muscle-specific association was observed with peripheral responses (M-max), supporting a stronger relationship between corticospinal drive and Hebbian plasticity in the distal than in the proximal arm muscle. Together, these findings highlight muscle-specific differences in the capacity for Hebbian plasticity and suggest that such differences may be important when developing targeted neuromodulation strategies for upper-limb rehabilitation.

### Hebbian plasticity across muscle groups

In line with previous studies, we found that Hebbian stimulation effectively enhanced corticospinal excitability in both the biceps brachii [17] and FDI [10, 18]. In the present study, we extended this work by directly comparing the effects of Hebbian stimulation between a proximal upper-limb muscle, the biceps brachii, and a distal hand muscle, the FDI. We found that MEP amplitude increased significantly more in the FDI than in the biceps, suggesting greater susceptibility to Hebbian plasticity in the distal upper limb. Our findings are consistent with emerging evidence that the magnitude of activity-dependent plasticity differs across upper-limb muscles. Even among proximal muscles, although MEP amplitude increased in both the triceps and biceps following Hebbian stimulation, the enhancement was significantly greater in the biceps, highlighting differential susceptibility to Hebbian plasticity across proximal upper-limb muscles [16]. Similarly, pairing TMS with electrical stimulation of motor points produced the largest increase in corticospinal excitability in intrinsic hand muscles, smaller increases in the flexor digitorum superficialis, and no detectable changes in the extensor digitorum communis [15]. Together, these findings suggest that the capacity for plasticity induced by neurostimulation may vary systematically across muscles, with distal muscles potentially showing greater responsiveness to activity-dependent stimulation.

Evidence from motor training paradigms further supports greater plasticity of distal upper-limb representations. Ballistic activation of distal upper-limb muscles during repetitive thumb-training tasks induces rapid cortical plasticity, resulting in changes in the direction of motor-evoked responses in the trained digit [19]. When ballistic training was compared between proximal and distal upper-limb joints, the largest training-related changes in corticospinal excitability were observed for muscles controlling distal joints, suggesting greater plasticity in distal than proximal upper-limb representations [20]. Similarly, it has been reported that cortical maps elicited by using TMS before and after visuomotor tracking training with the FDI and proximal (biceps brachii) muscle were larger in the FDI than the biceps brachii. Indeed, a previous study assessed the influence of elbow training on corticospinal excitability and found that repetitive elbow flexion causes only small increases in biceps MEPs [21]. Thus, consistent with our results, while both proximal and distal muscles retain substantial capacity for activity-dependent plasticity, our findings suggest that distal muscles may exhibit a greater response to Hebbian stimulation.

A critical question is why larger changes in MEP amplitude were observed in the FDI compared to biceps brachii. Anatomical and electrophysiological studies have demonstrated that corticospinal projections differ between proximal and distal upper-limb muscles. In non-human primates, distal muscles, particularly those of the hand, receive denser direct monosynaptic corticospinal–motoneuronal projections than proximal arm muscles, which receive less prominent monosynaptic corticospinal input [1, 2]. Consistent with these observations, studies in humans have shown stronger corticospinal facilitation of finger motoneurons than proximal arm motoneurons, as assessed using PSTHs of single motor unit discharges [3]. Likewise, MEPs elicited by transcranial magnetic stimulation are larger and have a lower threshold in finger muscles than in the biceps and triceps [4]. This is consistent with our results showing that the resting motor threshold was lower in the FDI than in the biceps brachii. This is also consistent with our findings showing that changes in MEP amplitude were associated with maximal MEP size in the FDI but not in the biceps, whereas no muscle-specific association was observed with peripheral responses (M-max), further supporting a stronger relationship between corticospinal drive and Hebbian plasticity in the distal than in the proximal arm muscle.

### Functional considerations

Notably, most previous neurostimulation studies targeting different upper-limb muscles have used similar stimulation parameters [22], leaving it unclear whether the capacity to induce plasticity varies across muscles. Our findings suggest that one-size-fits-all approaches may not fully account for differences in corticospinal organization. In particular, the smaller facilitation observed in the biceps brachii raises the possibility that proximal upper-limb muscles may require different stimulation parameters or complementary interventions to achieve levels of plasticity comparable to those observed in the FDI. This interpretation is consistent with evidence that Hebbian stimulation targeting lower-limb muscles required a greater number of paired stimuli than protocols targeting upper-limb muscles [10, 23]. These considerations are important because several studies have demonstrated that Hebbian stimulation can increase corticospinal excitability in targeted muscles in both neurologically intact individuals and people with spinal cord injury [10–13], and may also enhance motor recovery [11, 12]. Thus, optimizing stimulation parameters according to the muscle being targeted may be important for maximizing the therapeutic potential of Hebbian stimulation.

Our findings also highlight the challenges of inducing plasticity in the biceps brachii. Differences in the capacity for recovery between proximal and distal muscles are less likely to account for our findings, as evidence suggests that proximal muscles, including those of the upper arm, may demonstrate greater recovery after spinal cord injury than distal hand muscles, even when accounting for lesion distance [24]. Since the greater facilitation observed in the FDI was present in neurologically intact individuals, these suggest that these muscle-specific differences may reflect intrinsic properties of the intact motor system and that these differences need to be considered after neurological injury. Our results further suggest that intrinsic differences in corticospinal organization may contribute to the muscle-dependent capacity for Hebbian plasticity, consistent with the ability of our stimulation protocol to target corticospinal–motoneuronal connections. Understanding these differences may help guide the development of muscle-specific neuromodulation strategies for upper-limb rehabilitation. Future studies will be important to determine whether similar muscle-dependent effects are observed in clinical populations with altered corticospinal organization.

## Data Availability

All data produced in the present study are available upon reasonable request to the authors

